# Examining the impact of gestational diabetes genetic susceptibility variants on maternal glucose levels during and post-pregnancy

**DOI:** 10.1101/2025.06.26.25330189

**Authors:** Aminata H. Cissé, Alan Kuang, Catherine Allard, Justiina Ronkainen, Robin N. Beaumont, Sylvain Sebert, Denise M. Scholtens, Andrew T. Hattersley, Marja Vääräsmäki, Eero Kajantie, Luigi Bouchard, Patrice Perron, Elina Keikkala, Marie-France Hivert, William L. Lowe, Alice E. Hughes, Rachel M. Freathy

## Abstract

**Aim:** Gestational diabetes (GDM) has important environmental and genetic components. Genetic variants associated with GDM (n=14 SNPs) were recently classified into 2 groups: those with stronger effects on type 2 diabetes than GDM (Class-T, 3 SNPs) and those with stronger effects on GDM than on type 2 diabetes (Class-G, 8 SNPs), leaving 3 SNPs unclassified. It was suggested that the Class-G variants contribute to hyperglycaemia predominantly during gestation, but it is not known whether the effects of the two variant classes on maternal glucose levels vary with pregnancy status. We aimed to compare the effects of GDM-associated variants on glucose levels (fasting glucose and 2-hour post-OGTT) measured during vs. after pregnancy in longitudinal cohorts.

**Methods:** We calculated genetic scores (GS) by class (T_GS and G_GS) and overall (All_GS) in 10,225 pregnant women and 4,763 women post-pregnancy (mean 10.5 years post pregnancy) from 8 datasets representing 4 ancestrally-diverse cohorts: EFSOCH, Gen3G, HAPO and FinnGeDi. We used linear regression models adjusted for ancestry principal components to investigate associations between standardised GS and glucose levels during or after pregnancy. Analyses were performed separately in each dataset and then combined using inverse-variance weighted random-effects meta-analyses.

**Results:** In the meta-analysis, All_GS was associated with fasting glucose both during and after pregnancy (β[95%CI], in mmol/L per 1SD higher GS = 0.06 [0.04;0.08] during vs. 0.06 [0.04;0.07] post-pregnancy). All_GS was also associated with 2-hour post-OGTT glucose levels during pregnancy but not after (0.10 [0.04; 0.15] during vs. 0.01 [-0.04; 0.07] post-pregnancy). Both G_GS and T_GS showed consistent associations with fasting glucose during and post pregnancy (0.06 [0.04; 0.08] during and 0.05 [0.03; 0.07] post pregnancy for G_GS; 0.02 [0.01; 0.02] during and 0.02 [-0.001; 0.05] post pregnancy for T_GS). G_GS showed weak evidence of association with 2-hour glucose levels during pregnancy (0.06 [-0.002; 0.11]) and no association with 2-hour glucose levels post pregnancy (-0.03 [-0.08; 0.03]). However, T_GS was associated with 2-hour glucose during pregnancy and post pregnancy (0.10 [0.04; 0.16] and 0.06 [0.01; 0.12]).

**Conclusion:** Genetic scores for GDM have consistent associations with fasting glucose levels during and after pregnancy. This finding suggests that biological pathways underlying GDM genetic susceptibility to fasting hyperglycaemia are not pregnancy specific. However, the results for All_GS and 2-hour glucose provide evidence that some genetic associations with postprandial glucose may be stronger in pregnancy and should be followed up in larger samples.

## Introduction

Gestational diabetes mellitus (GDM), defined as glucose intolerance occurring or first observed during pregnancy, presents significant health risks for both mother and fetus [1]. Offspring of mothers who have had GDM are more likely to have higher birthweight and to develop obesity and metabolic disorders at an earlier age [2, 3]. In addition, GDM is a strong risk factor for developing type 2 diabetes in later life [4, 5]. Up to 50% of women who have had GDM develop prediabetes or type 2 diabetes within 10 to 15 years postpartum [6–8].

Genome-wide association studies (GWAS) have made progress in identifying genetic loci underlying variation in GDM risk. The GENetics of Diabetes In Pregnancy (GenDIP) Consortium performed a multi-ancestry meta-analysis of GWAS including 5,485 women with GDM and 347,856 controls, identifying five loci robustly associated with GDM [9]. Among these, four loci had previously been reported for type 2 diabetes, suggesting shared genetic pathways between the two conditions. Similarly, a study conducted in China on more than 30,000 pregnant women identified 4 loci associated with GDM, among which *MTNR1B* exhibited the strongest association with GDM while having a more modest effect on type 2 diabetes risk [10]. Kwak et al. also revealed that variants in *CDKAL1* and *MTNR1B* exhibited a stronger association with GDM than with type 2 diabetes in a two-stage genome-wide association analysis conducted among Korean women [11]. The largest GWAS of GDM to date, conducted by Elliott et al., included 12,332 women with GDM and 131,109 controls from Finland. This study assessed the shared genetic aetiology between GDM and type 2 diabetes [12]. They identified 13 loci for GDM, many of which also influence type 2 diabetes risk, and classified the loci into two categories. In one category, the variants had stronger effects on type 2 diabetes than GDM (Class-T), and in the other (Class-G), the variants had stronger effects on GDM than type 2 diabetes. A possible inference from these findings was that Class-G loci related to mechanisms of glucose regulation specific to pregnancy. However, several of these loci were already known to influence other commonly measured glycaemic traits in non-pregnant individuals without diabetes, and it is not known if their effects are altered by pregnancy status [11, 13].

To investigate the extent to which genetic susceptibility to GDM highlights glucose regulation mechanisms that are disrupted specifically during pregnancy, we aimed to compare the effects of groups of GDM-associated variants on fasting, and 2-hour post-OGTT glucose levels measured during vs. after pregnancy, across women of diverse ancestries. We performed a meta-analysis of eight datasets representing four cohorts: EFSOCH, Gen3G, HAPO, and FinnGeDi. We hypothesised that genetic scores based on Class-G variants would show stronger associations with glucose levels during pregnancy than after pregnancy.

## Materials and methods

### Study descriptions

We analysed data from 10,225 pregnant women and 4,763 women post-pregnancy across eight datasets representing four cohorts. The datasets included the Exeter Family Study of Childhood Health (EFSOCH), the Genetics of Glucose Regulation in Gestation and Growth (Gen3G) study, the Hyperglycaemia and Adverse Pregnancy Outcome (HAPO) study divided into five different ancestry groups based on genetic similarity (HAPO-European, HAPO-Afro-Caribbean, HAPO-East-Asian, HAPO-South-Asian and HAPO-Mexican-American), and the Finnish Gestational Diabetes (FinnGeDi) study.

The inclusion criteria required participants to have at least one glucose measurement during pregnancy and/or at least one glucose measurement post-pregnancy, along with available genome-wide genotype data. Women with pre-existing type 1 or type 2 diabetes were excluded from all analyses. Women with glucose levels indicative of GDM, measured prior to any therapeutic intervention, were included in analyses regardless of subsequent treatment in most cohorts (see cohort specific details in Supplementary Materials).

Details of genotyping and maternal glucose phenotype measurements for each cohort are provided in more detail in the supplementary material.

### Generating genetic scores

We used summary statistics from the GWAS of GDM published by Elliott et al [12] to calculate the genetic scores, based on their classification. The overall genetic score (All_GS) included all identified variants (14 SNPs including 3 unclassified SNPs), while T_GS encompassed variants with stronger effects on type 2 diabetes than GDM (3 SNPs), and G_GS encompassed variants with stronger effects on GDM than type 2 diabetes (8 SNPs). A genetic score for an individual is equal to the sum of the genotype dosages for each GDM risk allele of each SNP, weighted by the log odds ratio for GDM based on the GWAS summary statistics [12].

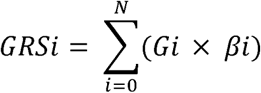

Where ***N*** is total number of variants included in the score, ***G_i_*** is dosage of risk alleles for SNP (a number between 0 and 2 based on the individual’s genotype) and β*_i_* is effect size (log odds ratio for GDM).

Each score was standardised within each cohort and ancestry group to a mean of 0 and SD of 1 for the regression analyses. A small number of SNPs were missing in different cohorts (*Supplementary Table S1*).

### Statistical analysis

Basic descriptive maternal characteristics were summarised for each dataset during and after pregnancy.

We used linear regression to test the associations between the standardised genetic scores and glucose levels (fasting or 2-hour glucose levels, in mmol/l) during and after pregnancy, in each dataset separately. The main model was adjusted for the first five principal components of ancestry in all cohorts, except Gen3G, which included the first four principal components; and the variables linked to the specificities of each cohort, such as genotyping batches, centre, etc. We referred to the “adjusted model” as the model with additional adjustment for maternal age at the time of glucose measurement. Then, we performed inverse-variance weighted random-effects meta-analyses to combine the results from the 8 datasets.

As a sensitivity analysis, we repeated the analyses including only women who had measurements both before and after pregnancy to test whether there was any difference in results when we compared the same women during and after pregnancy. We also conducted a leave-one-out meta-analysis to see if a given cohort is particularly different from the others.

In addition, we investigated two SNPs known to have particularly strong associations with type 2 diabetes and fasting glucose, respectively: the *TCF7L2* (rs34872471) SNP from Class-T and *MTNR1B* (rs10830963) SNP from Class-G have strong associations in non-pregnant individuals compared with other glycaemic trait SNPs [14] To evaluate their impact on the observed associations between the genetic scores and our outcomes, we conducted additional analyses by removing *TCF7L2* and *MTNR1B* from the construction of T_GS and G_GS, respectively.

## Results

The descriptive characteristics of the 8 cohorts (EFSOCH, Gen3G, HAPO-European, HAPO-Afro-Caribbean, HAPO-East-Asian, HAPO-South-Asian, HAPO-Mexican-American, and FinnGeDi) before and after pregnancy are presented in *Table 1*.

**Table 1:** Descriptions (mean (sd)) of the characteristics of each dataset.

|  | **EFSOCH** | **Gen3G** | **HAPO-AFR** | **HAPO-AMR** | **HAPO-EAS** | **HAPO-EUR** | **HAPO-SAS** | **FinnGeDi** |
| --- | --- | --- | --- | --- | --- | --- | --- | --- |
|  | **N=692** | **N=532** | **N=1372** | **N=965** | **N=2014** | **N=3235** | **N=129** | **N=1586** |
| **G_GS** | 2.74 (0.30) | 1.02 (0.30) | 0.93(0.19) | 1.91 (0.28) | 1.22 (0.32) | 1.94 (0.31) | 1.99 (0.33) | 2.83 (0.34) |
| **T_GS** | 0.67(0.13) | 0.68 (0.14) | 0.79 (0.14) | 0.66 (0.13) | 0.09 (0.09) | 0.67 (0.14) | 0.71 (0.14) | 0.66 (0.13) |
| **All_GS** | 3.82 (0.34) | 2.11 (0.33) | 2.13 (0.25) | 2.94 (0.32) | 1.73 (0.34) | 2.98 (0.34) | 3.09 (0.36) | 3.93 (0.37) |
| **Description of variables during pregnancy** |  |  |  |  |  |  |  |  |
| **Gestational age at glucose measurement (weeks)** | 28* | 26.5 (1.0) | 27.1 (1.7) | 27.0 (2) | 28.0 (1.6) | 28.4 (1.4) | 28.4 (1.2) | 24.6 (6.3) |
| **Maternal age at glucose measurement (years)** | 30.5 (5.2) | 28.9 (4.1) | 25.7 (5.7) | 28.9 (5.4) | 29.2 (5.5) | 31.2 (5.2) | 29.4 (4.7) | 30.9 (5.3) |
| **Fasting glucose (mmol/l)** | 4.3 (0.4) | 4.2 (0.3) | 4.5 (0.4) | 4.6 (0.4) | 4.4 (0.3) | 4.5 (0.4) | 4.7 (0.4) | 5.0 (0.6) |
| **2-h glucose (mmol/l)** | NA | 5.7 (1.2) | 6.0 (1.2) | 6.1 (1.3) | 6.5 (1.3) | 6.0 (1.2) | 6.5 (1.6) | 6.9 (1.7) |
| **Description of variables outside pregnancy** |  |  |  |  |  |  |  |  |
|  | **N=390** | **N=352** | **N=636** | **N=421** | **N=990** | **N=1580** | **N=119** | **N=275** |
| **Maternal age at glucose measurement (years)** | 36.7 (5.0) | 34.6 (4.1) | 38.0 (5.9) | 41.1 (5.5) | 41.7 (4.9) | 43.8 (4.9) | 41.2 (4.8) | 45.1 (5.2) |
| **Fasting glucose (mmol/l)** | 4.6 (0.4) | 4.7 (0.4) | 5.0 (0.8) | 5.2 (0.8) | 5.2 (0.8) | 5.1 (0.5) | 5.2 (0.6) | 5.5 (0.6) |
| **2-h glucose (mmol/l)** | NA | 5.2 (1.3) | 6.5 (2.2) | 6.6 (2) | 6.7 (2.3) | 6.0 (1.6) | 6.8 (2) | 6.0 (1.9) |

\* Fasting glucose during pregnancy in EFSOCH samples were taken at 28 weeks' gestation

### Genetic scores had consistent effects on fasting glucose levels during and after pregnancy

In our meta-analyses, both G_GS (8 SNPs) and T_GS (3 SNPs) showed positive associations with fasting glucose during and post pregnancy (β [95%CI] in mmol/L per 1 SD higher GS = 0.06 [0.04; 0.08] during and 0.05 [0.03; 0.07] post pregnancy for G_GS; 0.02 [0.01; 0.03] during and 0.02 [-0.0006; 0.05] post pregnancy for T_GS, *Figure 1*). All_GS (14 SNPs) was also associated with fasting glucose levels both during and post pregnancy with similar effects (0.06 [0.04;0.08] vs. 0.06 [0.04;0.07]). After adjustment for maternal age (*Supplementary Figure S1*), genetic scores were similarly associated with fasting glucose during pregnancy (0.06 [0.04; 0.09], 0.02 [0.01; 0.03] and 0.06 [0.04; 0.09] respectively for G_GS, T_GS and All_GS) and after pregnancy (0.06 [0.03; 0.07], 0.02 [0.0004; 0.05] and 0.06 [0.04;0.07] respectively for G_GS, T_GS and All_GS). Heterogeneity between studies was high for the association estimates during pregnancy (maximum I^2^ = 86.6% (p< 0.0001).

**Figure 1:**
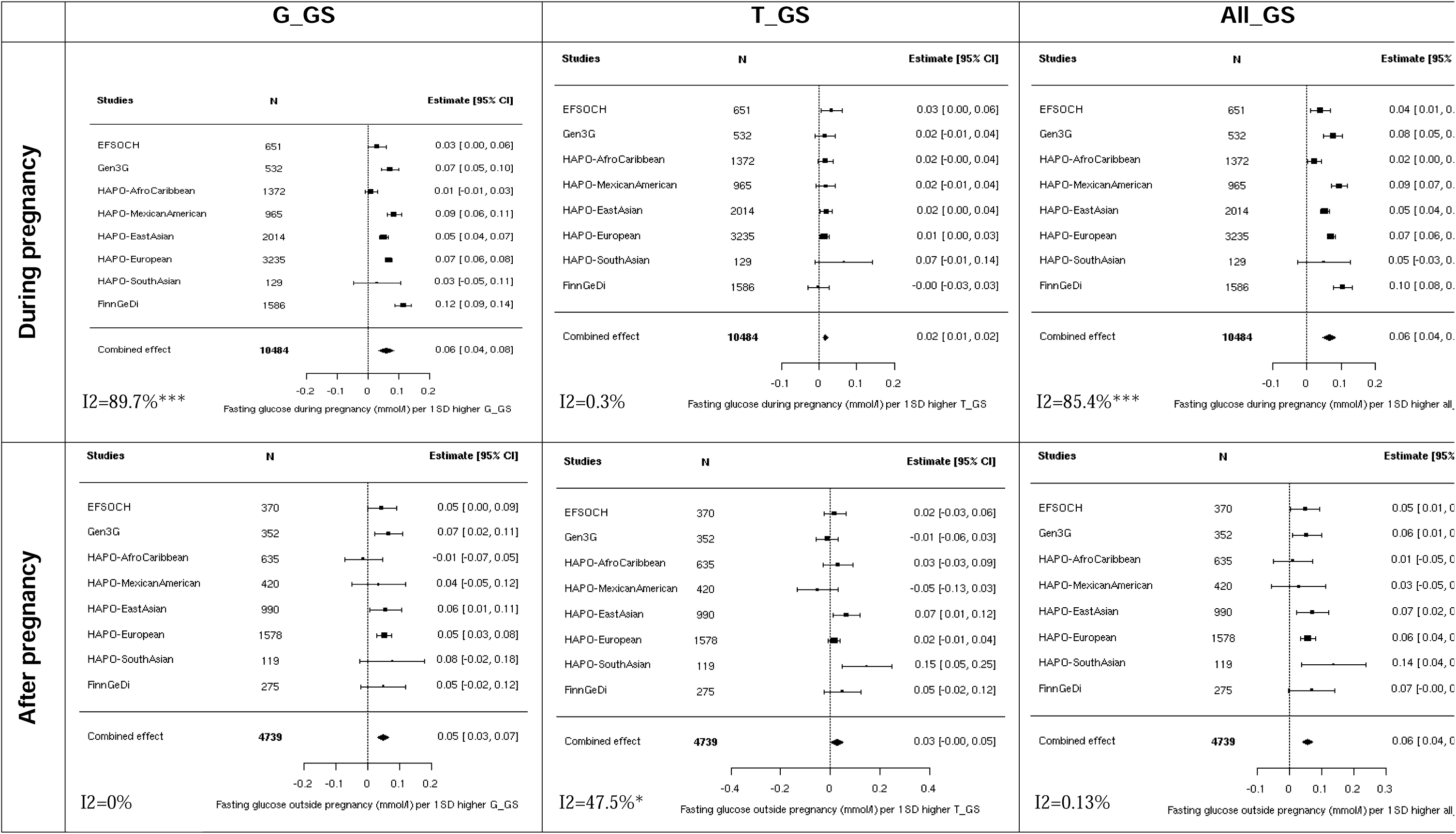
Meta-analysis of the association between fasting glucose and genetic scores. Analyses adjusted solely for principal components and cohort-specific variables (to address unique characteristics of each data. Heterogeneity statistics (I^2^) are included in the bottom left of each plot. G_GS = 8 SNPs; T_GS = 3 SNPs; All_GS = 13 SNPs. ***: p-value <.0001; **: p-value < 0.001; *: p-value < 0.05

### Associations with 2-hour glucose levels varied according to genetic scores, with weaker evidence observed after pregnancy

G_GS showed weak evidence of association with 2-hour glucose levels during pregnancy (0.06 [-0.002; 0.11]) and no association with 2-hour glucose levels post pregnancy (-0.03 [-0.08; 0.03], p > 0.05, *Figure 2*). In contrast, T_GS was associated with 2-hour glucose levels both during and after pregnancy (0.10 [0.04; 0.16] and 0.06 [0.01; 0.12], respectively). All_GS was associated with 2-hour glucose levels during pregnancy (0.10 [0.04; 0.15]) but not afterward (0.01 [-0.04; 0.07]). After adjusting for maternal age, the results remained consistent: 0.06 [-0.002; 0.11] during pregnancy and -0.03 [-0.08; 0.03] post pregnancy for G_GS; 0.11 [0.04; 0.17] during pregnancy and 0.06 [0.01; 0.12] post pregnancy for T_GS; and 0.10 [0.04; 0.16] during pregnancy and 0.01 [-0.05; 0.07] post pregnancy for All_GS (*Supplementary Figure S2*). Heterogeneity between studies was high for the association estimates during pregnancy (maximum I^2^ = 78.2%; p= 0.001).

**Figure 2:**
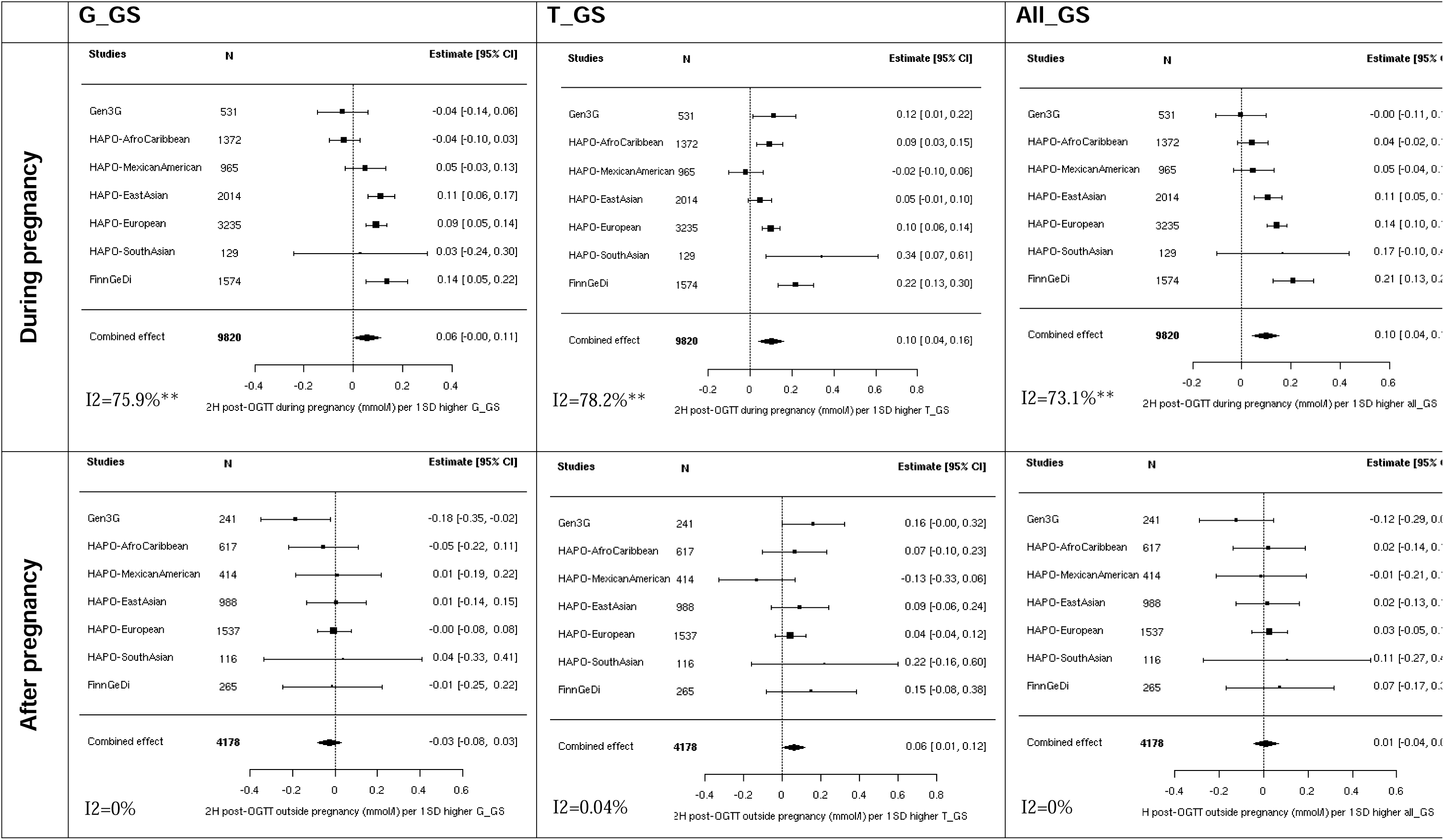
Meta-analysis of the association between 2-hour glucose and genetic scores. Analyses adjusted solely for principal components and cohort-specific variables (to address unique characteristics of each datas

### Sensitivity analyses showed consistent results with the main analyses

The results of the analyses including only women who had had measurements before and after pregnancy were consistent with the results of the main analysis, with wider confidence intervals due to the smaller sample size (*Supplementary Figures S3-S6*).

Leave-one-out sensitivity analyses showed that omitting HAPO–Mexican-American strengthened the G-GRS association between the G-GRS and 2-hour glucose during pregnancy and was the only instance that eliminated heterogeneity.

After removing the *MTNR1B* SNP rs10830963 from the G_GS calculation, associations between G_GS (now containing 7 SNPs) and fasting glucose in the meta-analysis remained, (0.04 [0.02; 0.07] during pregnancy and 0.03 [0.01; 0.04] post pregnancy, *Supplementary Figure S7*). The associations with 2-hour glucose also showed similar patterns to the main analysis, both during and after pregnancy, 0.002 [-0.04; 0.04] during pregnancy and -0.05 [-0.10; 0.01] post pregnancy, p > 0.05). After removing *TCF7L2* rs34872471 from the T_GS there was still evidence of association with 2-hour glucose during pregnancy (*Supplementary Figure S8*). However, removal of one SNP from a T_GS (now containing 2 SNPs) had a large impact on the score’s standard deviation, so it was not possible to make meaningful comparisons between effect sizes in this and the main analysis.

## Discussion

In this meta-analysis of up to 10,225 women from eight international, multi-ancestral cohorts, we have shown that the combined effects of gestational diabetes susceptibility variants on fasting glucose levels were similar during vs. after pregnancy. Each of the three genetic scores tested (G_GS: 8 variants with stronger effects on GDM than type 2 diabetes; T_GS: 3 variants with stronger effects on type 2 diabetes than GDM; and All_GS: 14 variants combining G_ and T_GS with 2 additional unclassified variants) was associated similarly with fasting glucose during and post-pregnancy. These findings do not align with our initial hypothesis that G_GS variants would exhibit stronger effects on glucose levels during pregnancy. However, we observed associations with 2-hour glucose that were stronger during pregnancy than post-pregnancy. The All_GS was associated with 2-hour glucose levels during pregnancy but not post-pregnancy, and the 95% confidence intervals around the post-pregnancy estimate did not include the during-pregnancy estimate (and vice versa), which suggests an overall difference in the combined effect estimate. This did not appear to be driven by the T_GS, which was similarly associated with 2-hour glucose levels during and after pregnancy, but by the weaker post-pregnancy G_GS associations.

In the most recent GWAS of GDM from Elliott et al., it was suggested that certain SNPs (primarily those in Class-G) may influence glucose homeostasis predominantly during pregnancy [15]. Our findings did not demonstrate differential effects of the genetic scores on fasting glucose levels during vs. after pregnancy. Variants at the majority of the GDM-associated loci have previously been associated with fasting glucose at genome-wide significance outside of pregnancy, including five out of the eight Class-G loci, two out of the three Class-T loci and two out of the three unclassified loci [12]. The Class-G variants include *MTNR1B* rs10830962 and *G6PC2* rs1402837, which are both strongly and consistently associated with fasting glucose outside of pregnancy [16]. In line with this, a recent analysis of individual SNP effects on fasting glucose in the HAPO study (which contributed to our meta-analysis) found consistent associations for SNPs near *MTNR1B* (rs10830962) and *G6PC2* (rs560887, in LD with rs1402837 (r2=0.1 in Europeans)) during pregnancy and 11-14 years postpartum [16]. However, a *PCSK1* variant (rs12332295, in LD with the G_GS variant rs1820176; r2=0.4 to 1.0 across multiple ancestries) primarily influenced fasting glucose during pregnancy [17]. It is possible then that while many of the Class-G variants have consistent effects on fasting glucose during and outside pregnancy, some may have more pronounced effects in gestation. Larger samples are needed for sufficient statistical power to interrogate these individual SNP effects.

We observed differences in the associations with 2-hour glucose levels across the G_GS and All_GS genetic scores, showing weaker evidence of association after pregnancy. Similarly, the recent analysis of individual SNPs in HAPO examined the differential effects of genetic variants on 2-hour glucose levels post-OGTT during and after pregnancy. Their results showed that the *MTNR1B* variant rs10830962 had a significant impact on 2-hour glucose levels during pregnancy that was not present post-pregnancy [17]. Notably, neither this variant nor the *G6PC2* rs1402837 fasting glucose variant have been robustly associated with 2-hour post-OGTT glucose levels in large GWAS meta-analyses of non-pregnant individuals [16]. Our results suggest a possible modulation of postprandial glucose regulation during pregnancy, with limited influence on fasting glucose levels. The hormonal and metabolic changes that occur during pregnancy, could specifically impact postprandial glucose metabolism by enhancing the effect of certain genetic variants [17]. However, the influence on fasting glucose may remain more stable, possibly due to different regulatory mechanisms involved in fasting glucose control.

The timing of glucose measurements during pregnancy may represent an important factor to consider in future studies. In the studies contributing to our meta-analysis, glucose levels were measured towards the end of the second and beginning of the third trimester, but results may have been different if measurements were taken earlier, as is sometimes done for high-risk pregnancies in some healthcare settings. Physiologically, pregnancy is associated with a progressive increase in insulin resistance due to hormonal changes, as well as relative β-cell dysfunction, particularly during the second and third trimesters [18–21]. These physiological changes could lead to a decrease in fasting glucose and an increase in postprandial glucose, in a normoglycemic pregnant women [18–21]. These metabolic adaptations are unique to pregnancy and may alter the expression of genetic effects over time. Some genetic variants may exert consistent effects regardless of gestational stage, while others might be more strongly associated with glucose levels in late pregnancy due to interactions with the evolving hormonal environment.

The differences in diagnostic criteria between GDM and type 2 diabetes may be relevant to the aetiology of the GDM variant classifications and our results. GDM diagnosis is characterised by a lower fasting plasma glucose threshold than type 2 diabetes (5.1 mmol/L vs 7.0 mmol/L) [22], reflecting the importance of maternal fasting glycaemia on risk of adverse pregnancy outcomes [23]. In contrast, although type 2 diabetes is less frequently diagnosed by OGTT than GDM, studies have shown that most individuals diagnosed with type 2 diabetes will have a raised 2-hour glucose ≥11.1 mmol/L [25, 26]. We found that the G_GS was more consistently associated with fasting glycaemia, whereas the T_GS showed more consistent associations with 2-hour glucose levels. This may, in part, relate to the differences in diagnostic criteria for GDM and type 2 diabetes impacting on the classification of GDM susceptibility variants.

The strengths of our work are the inclusion of diverse ancestry groups and the large, combined sample size. However, there were also limitations to our work. We noticed a high heterogeneity between studies, suggesting unaccounted differences not addressed here, even though sensitivity analyses confirmed the robustness of the associations. Some SNPs included in the generation of the genetic score were frequently missing across cohorts. We hypothesised that some of the heterogeneity may have arisen due to differences in standard deviations of genetic scores leading to inconsistent estimates of effect sizes once the scores were standardized, but further “leave-one-out” analyses suggested these differences did not account for much of the heterogeneity (*Supplementary Table S2*). Heterogeneity between studies may also have resulted from the variable cohort compositions, for example, FinnGeDi is a GDM case-control study, so was enriched for women at high risk, in contrast to the more general population cohorts of pregnant women from the other studies. Additionally, the summary statistics used in this work were derived from GWAS analyses conducted in the Finnish population [15], which may limit the applicability of results to other European or non-European cohorts. Finally, the limited number of SNPs in the genetic scores (especially Class-T) may have made them vulnerable to influences by individual SNPs with strong effects. For example, the GDM-susceptibility variants at *MTNR1B* (included in G_GS) and *TCF7L2* (included in T_GS) have relatively large effects compared with variants in the same groups. However, our sensitivity analyses showed that neither SNP was solely responsible for driving the observed associations with glucose levels because the associations were evident with these SNPs removed.

To conclude, genetic scores associated with GDM have consistent effects on fasting glucose levels during and after pregnancy. This finding reflects the importance of the fasting glucose threshold in GDM diagnosis and indicates biological pathways underlying GDM genetic susceptibility that are not pregnancy specific. However, the associations between predominantly GDM-associated variants and 2-hour glucose levels suggest genetic associations with postprandial glucose may differ in pregnancy. The observed heterogeneity highlights the need to explore factors that may vary between studies or populations.

## Funding and acknowledgements

The Exeter Family Study of Childhood Health (EFSOCH) was supported by□Southwest□NHS Research and Development, Exeter NHS Research and Development, the Darlington Trust and the National Institute for Health and Care Research Exeter Clinical Research Facility. This work was also supported by the National Institute for Health and Care Research Exeter Biomedical Research Centre. The views expressed are those of the authors and not necessarily those of the NIHR or the Department of Health and Social Care. Genotyping of the EFSOCH study samples was funded by□the□Wellcome□Trust and Royal Society grant□104150/Z/14/Z.

Gen3G was funded by the Fonds de recherche du Québec—Santé (FRQS) operating grant (to MIZIFH, grant #20697); the Canadian Institute of Health Research (CIHR) operating grants (to MIZIFH grant #MOP 115071 and to LB #PJT152989), Diabète Québec, Internal funding support from le Centre de recherche du CHUS and l’Université de Sherbrooke, and from American Diabetes Association (to MFH #1-15-ACE-26).

The HAPO Study and HAPO ancillary studies that provided genetics data were funded by the National Institutes of Health (grants DK095963, DK117491, HD34242, HD34243, HG-004415).

The FinnGeDi study has been funded by the French National Research Agency (Agence Nationale de la Recherche), ANR-16-CE17-0017-01; Fondation Francophone pour la Recherche sur le Diabète (FFRD), which is sponsored by Fédération Française des Diabètiques (FFD), Abbott, AstraZeneca, Eli Lilly, Merck Sharp & Dohme (MSD) and Novo Nordisk. This work was also supported by the Agence Nationale de la Recherche (ANR) grants European Genomic Institute for Diabetes (E.G.I.D), ANR-10-LABX-0046, a French State fund managed by ANR under the frame program Investissements d′Avenir I-SITE ULNE / ANR-16-IDEX-0004 ULNE and from the National Center for Precision Diabetic Medicine – PreciDIAB, which is jointly supported by the French National Agency for Research (ANR-18-IBHU-0001), by the European Union (FEDER), by the Hauts-de-France Regional Council and by the European Metropolis of Lille (MEL); The Research Council of Finland (project grant and to EK grant #330343); Diabetes Research Foundation; Foundation for Pediatric Research; Juho Vainio Foundation; Novo Nordisk Foundation; Signe and Ane Gyllenberg Foundation; Sigrid Jusélius Foundation; Yrjö Jahnsson Foundation for Pediatric Research; Finnish Medical Foundation; Research Funds of Oulu University Hospital and Helsinki University Hospital (state grants); Päivikki and Sakari Sohlberg Foundation; Suorsa Foundation; Otto A Malm Foundation and Finnish Foundation for Cardiovascular Research.

## Authors acknowledgements/fundings

A.H.C., R.N.B. and R.M.F. were supported by a Wellcome Senior Research Fellowship (WT220390). J.R. was supported by European Union’s Horizon 2020 (grant number 874739, LongITools).

J.R. was supported by European Union’s Horizon 2020 (grant number 874739, LongITools).

A.E.H. was supported by a National Institute of Health Research (NIHR) Academic Clinical Fellowship.

## Author’s contributions

A.H.C. and R.M.F. conceptualized the study, developed the methodology, and wrote the initial draft of the manuscript. A.H.C., A.K., C.A., and J.R. conducted the statistical analyses. R.N.B., D.M.S., El.K. and A.E.H. supported analyses and were responsible for data curation and validation. S.S., A.T.H., M.V., Ee.K., L.B., P.P., M.F.H., and W.L.L. contributed to data collection and provided essential resources for the study. All authors interpreted data, reviewed the manuscript drafts, provided critical feedback, approved the final version, and are accountable for all aspects of the work.

## Data Availability Statement

To request access to the EFSOCH (Exeter Family Study of Childhood Health) dataset, researchers should contact the Exeter Clinical Research Facility (CRF) via email at.

HAPO phenotype data are currently available through dbGaP (www.ncbi.nlm.nih.gov/gap). The genotypic information is available from on reasonable request.

Gen3G data used in this study may be obtained by contacting M-FH and PP.

Request FinnGeDi data availability by contacting MV.

