## Supplementary material for "Examining the impact of gestational diabetes genetic susceptibility variants on maternal glucose levels during and post-pregnancy"

### Supplementary Information

Contents:

1. Information about phenotype and genotype preparation in contributing studies
2. Supplementary Tables (S1, S2) and Figures (S1-S8)
3. References

#### Information about phenotype and genotype preparation in contributing studies

**EFSOCH:** The Exeter Family Study of Childhood Health recruited a total of 986 healthy babies after exclusions, between 1999-2004. A full description of this study was published previously [1]. In this study, fasting blood samples were collected from the mothers during the 28th week of gestation in all women, after a 10-hour overnight fast. The samples were taken in the morning at the parents' homes. Plasma glucose concentrations were subsequently measured at the pathology laboratories of the Royal Devon and Exeter Hospital, using standard reagents provided by the manufacturer and analysed on Modular systems [1, 2]. The same protocol was followed at a median of 5.14 years post-pregnancy, when all women were invited for a post-pregnancy study visit, and a total of 523 women took part [3]. The 2-hour glucose levels post-OGTT were not measured in the EFSOCH study, either during pregnancy or after pregnancy, so EFSOCH only contributed to fasting glucose analyses in the current study.

Genotyping for the entire sample was performed using the Illumina HumanCoreExome-24 array. Raw genotyping data underwent quality control to ensure reliability, with exclusion criteria including a genotype call rate below 95%, SNPs deviating significantly from Hardy-Weinberg equilibrium ($p < 1 \times 10^{-6}$). Additionally, participants' phenotypic sex and familial relationships were validated using genotype data analysed with the KING software [4]. Imputation was conducted using the TopMed reference panel via the Michigan Imputation Server. Only SNPs with an imputation quality score above 0.4 and a minor allele frequency exceeding 0.1% were retained for analysis.

For this analysis, the EFSOCH sample included 692 pregnant women and 390 post pregnancy women with genotype and phenotype data available.

**Gen3G:** The Genetics of Glucose Regulation in Gestation and Growth (Gen3G) study is a population-based cohort from Quebec, Canada, which recruited 1,024 pregnant women without diabetes between January 2010 and June 2013. Its primary aim is to enhance understanding of the biological, environmental, and genetic factors influencing glucose regulation during pregnancy and their impact on fetal and offspring development. The details of the cohort description have been published elsewhere [5]. In the Gen3G cohort, glycemia was measured between 24 and 30 weeks' gestation at fasting and 2-hour post 75 g-OGTT using the hexokinase method (Roche Diagnostics, Indianapolis, IN) [6, 7]. At 3 and 5 years after delivery, Gen3G participants were invited for a follow-up visiting including collection of fasting blood and a subgroup of women also completed a 75g-OGTT at the 5-year visit with blood samples at fasting and 2h; plasma glucose was measured with the same laboratory methods as for pregnancy samples [8].

Maternal DNA was extracted from blood buffy coats using the Gentra Puregene Blood Kit (Qiagen, Mississauga, Canada) [6]. Genomic data were generated with the Expanded Multi-Ethnic Global Array (Illumina). SNPs included in the analysis had a call rate above 95% and adhered to Hardy-Weinberg equilibrium ($p>1\times10^{-6}$). To ensure data accuracy, SNPs were cross-checked for inconsistencies between mother-child pairs or multiple pregnancies from the same individual, and biologically implausible samples were excluded. Imputations were carried out using eagle for phasing, the TOPMed R2 reference panel on TOPMed imputation server using default parameter. Only SNPs with a R2>0.8 were used in the analyses.

For this analysis, only women with complete genomic data and at least one glucose level was included (for the current analyses, 35 women receiving treatment for GDM were excluded), corresponding to 532 women during pregnancy and 352 women outside pregnancy for a total of 533 participants.

**HAPO:** The Hyperglycemia and Adverse Pregnancy Outcome (HAPO) Study is a prospective observational study involving the recruitment of around 25,000 pregnant women across nine countries. The study's details have been documented in prior publications [9, 10]. In the HAPO study, women underwent 75g OGTT between 24 and 32 weeks of pregnancy. Blood samples were collected for fasting glucose analysis, as well as for postprandial glucose measurements at 2 hours post-OGTT. At approximately 10-14 years post pregnancy, participants in the HAPO study were selected according to inclusion criteria: term birth (gestational age ≥ 37 weeks), absence of major neonatal malformations or foetal/neonatal death, caregivers and participants blinded to the initial HAPO study results. Fasting and 2-hour blood glucose (75g OGTT) were measured by the hexokinase method on a Beckman-Coulter SYNCHRON LX analyser in plasma.

The genotyping for the HAPO cohort involved comprehensive data collection across multiple ethnic groups, including European, African Caribbean, East Asian, South Asian, and Mexican American populations. Genomic DNA was extracted from maternal blood samples, and genotyping was performed using platforms such as the Illumina HumanOmni1-Quad BeadChip, or similar, depending on the specific cohort. SNPs had call rates greater than 95%, and their alignment with Hardy-Weinberg equilibrium was confirmed. Imputation was carried out against the TOPMed reference panel on the TOPMed imputation server. The detailed methodology of genotyping in the HAPO study, including how these specific ethnic analyses were carried out, has been published elsewhere [11–16].

In our study, analyses were performed separately for each HAPO subgroup. These subgroups consisted of 1372, 965, 2014, 3235, and 129 women during pregnancy; and 636, 421, 990, 1580, and 119 women post pregnancy, representing African Caribbean, Mexican American, East Asian, European, and South Asian ancestries, respectively.

**FinnGeDi :** The Finnish Gestational Diabetes Study (FinnGeDi) is a case-control study conducted within the Finnish population to investigate genetic and environmental factors associated with the development of gestational diabetes. Previously described in detail, this study recruited a total of 1,066 non-diabetic pregnant women and 1,146 women diagnosed with gestational diabetes, between 2009 and 2012 [17, 18]. Blood samples were taken to measure fasting glucose levels, as well as glucose levels at 2 hours following 75g OGTT, typically conducted between 24 and 28 weeks of gestation according to the Finnish National current Care Guidelines. Glucose data were collected both during pregnancy and consistently 10-15 years post pregnancy.

The FinnGeDi genetic data were divided into two datasets because they were genotyped at different times. Maternal DNA was extracted from venous blood samples. Genotyping was conducted using Illumina Infinium Omni 2.5-8 for a subset of 516 samples and Illumina GSA 500k for a subset of 1455 samples. After quality control, variants with SNP-based call rate > 95% or Hardy-Weinberg equilibrium p > 0.0001 were included. Datasets were imputed separately using Finnish ancestry reference-panel SISu v3 (protocol: https://www.protocols.io/view/genotype-imputation-workflow-v3-0-e6nvw78dlmkj/v2). Eagle v2.3.5 was used for the genotype phasing and Beagle v4.1 was used for the imputation.

GS calculations were performed separately for each genotyping datasets and then combined in the regression analyses. A total of 1586 women with fasting, and 2-hour glucose data, along with genotype information during pregnancy, were included in our analysis. After pregnancy, 275 women had fasting and 2-hour glucose measurements.

#### Supplementary Tables and Figures

Supplementary Table S1: Availability of SNPs across cohorts

| Chromosome | Position, hg38 | rsid | Effect allele | other allele | Nearest gene | Class | Effect size (SE) | EFSOCH | Gen3G | HAPO-AFR | HAPO-AMR | HAPO-EAS | HAPO-EUR | HAPO-SAS | FinnGeDi | FinnGeDi |
|---|---|---|---|---|---|---|---|---|---|---|---|---|---|---|---|---|
| 2 | 27519736 | Rs780093 | C | T | GCKR | G | 0.12 (0.01) | ✓ | ✓ | ✓ | ✓ | ✓ | ✓ | ✓ | ✓ | ✓ |
| 2 | 168900844 | Rs1402837 | T | C | SPC25-G6PC2 | G | 0.11 (0.02) | ✓ | ✓ | ✓ | ✓ | ✓ | ✓ | ✓ | ✓ | ✓ |
| 3 | 123376465 | Rs6798189 | G | A | ADCY5 | Unclassified | 0.1 (0.02) | ✓ | ✓ | ✓ | ✓ | ✓ | ✓ | ✓ | ✓ | ✓ |
| 5 | 96360881 | Rs1820176 | T | C | PCSK1 | G | 0.14 (0.01) | ✓ | ✓ | ✓ | ✓ | ✓ | ✓ | ✓ | ✓ | ✓ |
| 6 | 20676183 | Rs34499031 | TAA | T | CDKAL1 | T | 0.12 (0.01) | ✓ | ✓ | ✓ | ✓ | ✓ | ✓ | ✓ | ✓ | ✓ |
| 6 | 151805650 | Rs537224022 | C | G | ESR1 | G | 0.45 (0.08) | ✓ | NA | NA | ✓ | NA | ✓ | ✓ | ✓ | ✓ |
| 9 | 22136490 | Rs1333051 | A | T | CDKN2B | Unclassified | 0.13 (0.02) | ✓ | ✓ | ✓ | ✓ | ✓ | ✓ | ✓ | ✓ | ✓ |
| 9 | 22134303 | Rs7019437 | G | C | CDKN2B | Unclassified | 0.04 (0.01) | ✓ | ✓ | NA | NA | NA | NA | NA | ✓ | ✓ |
| 10 | 112994312 | Rs34872471 | C | T | TCF7L2 | T | 0.17 (0.02) | ✓ | ✓ | ✓ | ✓ | ✓ | ✓ | ✓ | ✓ | ✓ |
| 11 | 92975544 | Rs10830963 | G | C | MTNR1B | G | 0.4 (0.01) | ✓ | ✓ | ✓ | ✓ | NA | ✓ | ✓ | ✓ | ✓ |
| 12 | 4275678 | Rs76895963 | T | G | CCND2 | T | 0.26 (0.04) | ✓ | ✓ | ✓ | ✓ | ✓ | ✓ | ✓ | ✓ | ✓ |
| 12 | 97457224 | Rs74628648 | C | T | NEDD1 | G | 0.17 (0.03) | ✓ | ✓ | ✓ | ✓ | ✓ | ✓ | ✓ | ✓ | ✓ |
| 16 | 81488676 | Rs2926003 | C | T | CMIP | G | 0.08 (0.02) | ✓ | ✓ | ✓ | ✓ | ✓ | ✓ | ✓ | ✓ | ✓ |
| X | 19380197 | Rs56381411 | C | T | MAP3K15 | G | 0.4 (0.06) | ✓ | NA | NA | NA | NA | NA | NA | ✓ | ✓ |

✓ = SNP available in the cohort NA = SNP missing in the cohort

Supplementary Table S2: Leave-One-Out study using the main analyses model, Impact of Cohort Exclusion on Heterogeneity ($I^2$)

| | | Fasting glucose during pregnancy | | Fasting glucose after pregnancy | | 2-hour glucose during pregnancy | | 2-hour glucose after pregnancy | |
|---|---|---|---|---|---|---|---|---|---|
| GS | ExcludedStudy | I2 | Pvalue_I2 | I2 | Pvalue_I2 | I2 | Pvalue_I2 | I2 | Pvalue_I2 |
| G_GS | EFSOCH | 90.49 | 1.07E-08 | 0 | 0.49 | NA | NA | NA | NA |
| | HAPO-Afro-Caribbean | 91.50 | 2.47E-09 | 0 | 0.56 | 73.06 | 0.004 | 0 | 0.994 |
| | HAPO-Mexican-American | 83.40 | 2.27E-04 | 0 | 0.99 | 53.39 | 0.093 | 0 | 0.489 |
| | HAPO-East-Asian | 90.52 | 1.62E-08 | 0 | 0.50 | 79.89 | 0.001 | 1.17 | 0.499 |
| | HAPO-European | 89.48 | 2.95E-09 | 0 | 0.50 | 73.46 | 0.002 | 8.41 | 0.508 |
| | HAPO-South-Asian | 88.82 | 7.24E-09 | 0 | 0.51 | 73.20 | 0.002 | 2.17 | 0.556 |
| | Gen3G | 91.41 | 2.01E-09 | 0 | 0.53 | 79.03 | 0.001 | 0 | 0.491 |
| | FinnGeDi | 85.25 | 3.61E-06 | 0 | 0.49 | 74.67 | 0.002 | 0 | 0.477 |
| T_GS | EFSOCH | 3.16 | 0.737 | 63.09 | 0.02 | NA | NA | NA | NA |
| | HAPO-Afro-Caribbean | 0.34 | 0.568 | 39.35 | 0.06 | 83.83 | 0.001 | 0.01 | 0.402 |
| | HAPO-Mexican-American | 0.58 | 0.568 | 62.34 | 0.02 | 82.71 | 0.001 | 10.68 | 0.242 |
| | HAPO-East-Asian | 0.16 | 0.570 | 43.32 | 0.07 | 67.81 | 0.016 | 0 | 0.752 |
| | HAPO-European | 3.30 | 0.585 | 0.36 | 0.07 | 79.60 | 0.002 | 6.44 | 0.255 |
| | HAPO-South-Asian | 0.02 | 0.600 | 58.15 | 0.03 | 79.85 | 0.001 | 19.51 | 0.274 |
| | Gen3G | 0 | 0.786 | 0.02 | 0.20 | 78.33 | 0.003 | 0.03 | 0.300 |
| | FinnGeDi | 0.01 | 0.813 | 57.08 | 0.03 | 51.06 | 0.033 | 0.03 | 0.294 |
| all_GS | EFSOCH | 86.64 | 2.24E-06 | 0.20 | 0.49 | NA | NA | NA | NA |
| | HAPO-Afro-Caribbean | 87.73 | 1.15E-06 | 0.18 | 0.48 | 69.93 | 0.013 | 0 | 0.993 |
| | HAPO-Mexican-American | 75.93 | 0.002 | 1.36 | 0.77 | 72.92 | 0.013 | 0 | 0.685 |
| | HAPO-East-Asian | 84.83 | 1.81E-05 | 0.11 | 0.52 | 76.16 | 0.005 | 0 | 0.688 |
| | HAPO-European | 84.79 | 2.46E-06 | 0.50 | 0.52 | 76.55 | 0.002 | 0 | 0.682 |
| | HAPO-South-Asian | 84.36 | 2.33E-06 | 0.0001 | 0.48 | 70.36 | 0.010 | 0 | 0.731 |
| | Gen3G | 87.80 | 7.42E-07 | 0 | 0.81 | 77.43 | 0.002 | 0 | 0.719 |
| | FinnGeDi | 82.49 | 4.97E-05 | 0.677 | 0.49 | 61.03 | 0.025 | 0 | 0.723 |

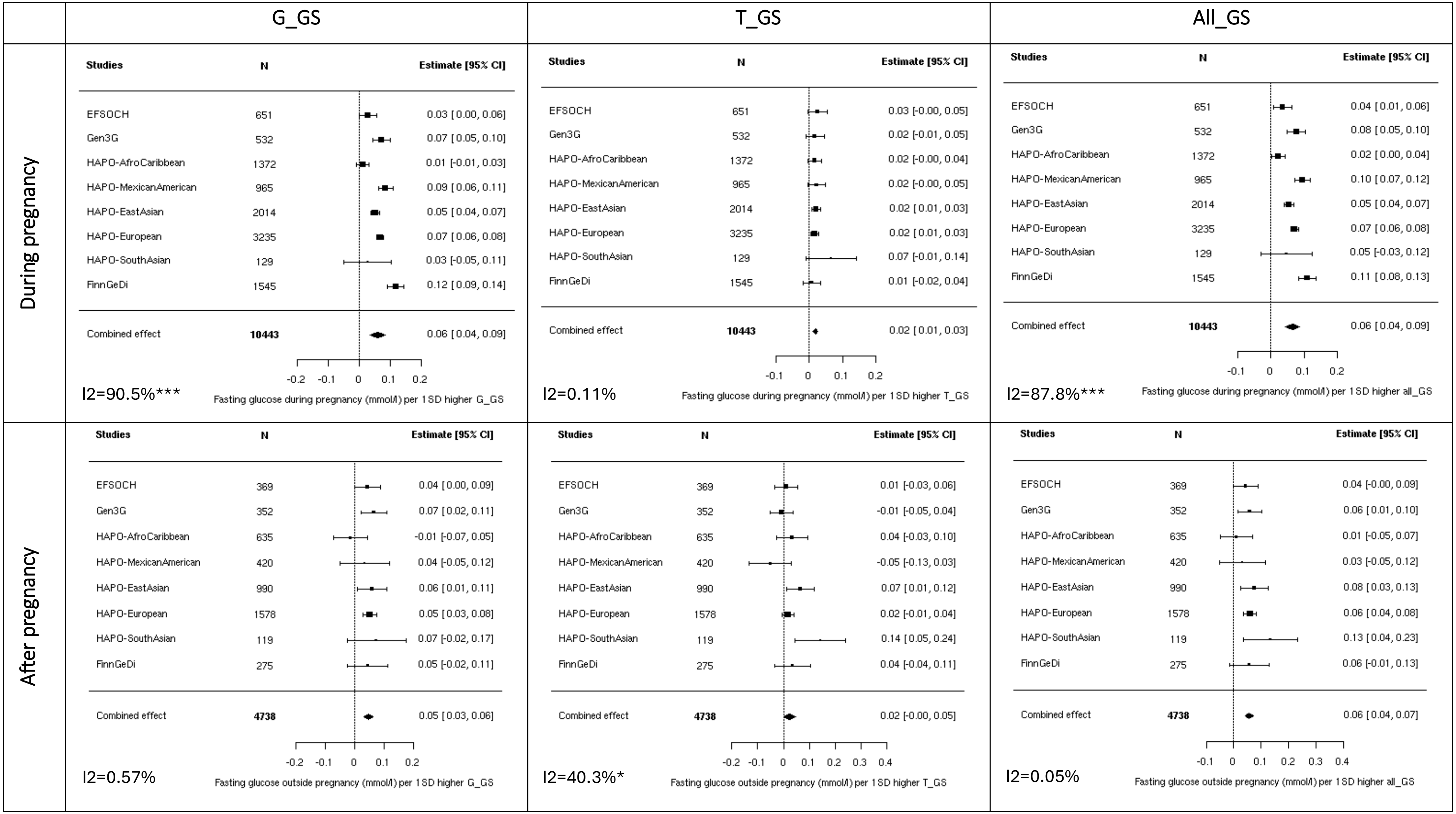


**Supplementary figure S1: Meta-analysis of the association between fasting glucose and genetic scores, adjusted for maternal age at glucose measurement**

Analyses also adjusted for principal components and cohort-specific variables (to address unique characteristics of each dataset). Heterogeneity statistics ($I^2$) are included in the bottom left of each plot.***: p-value <.0001; **: p-value < 0.001; *: p-value < 0.05

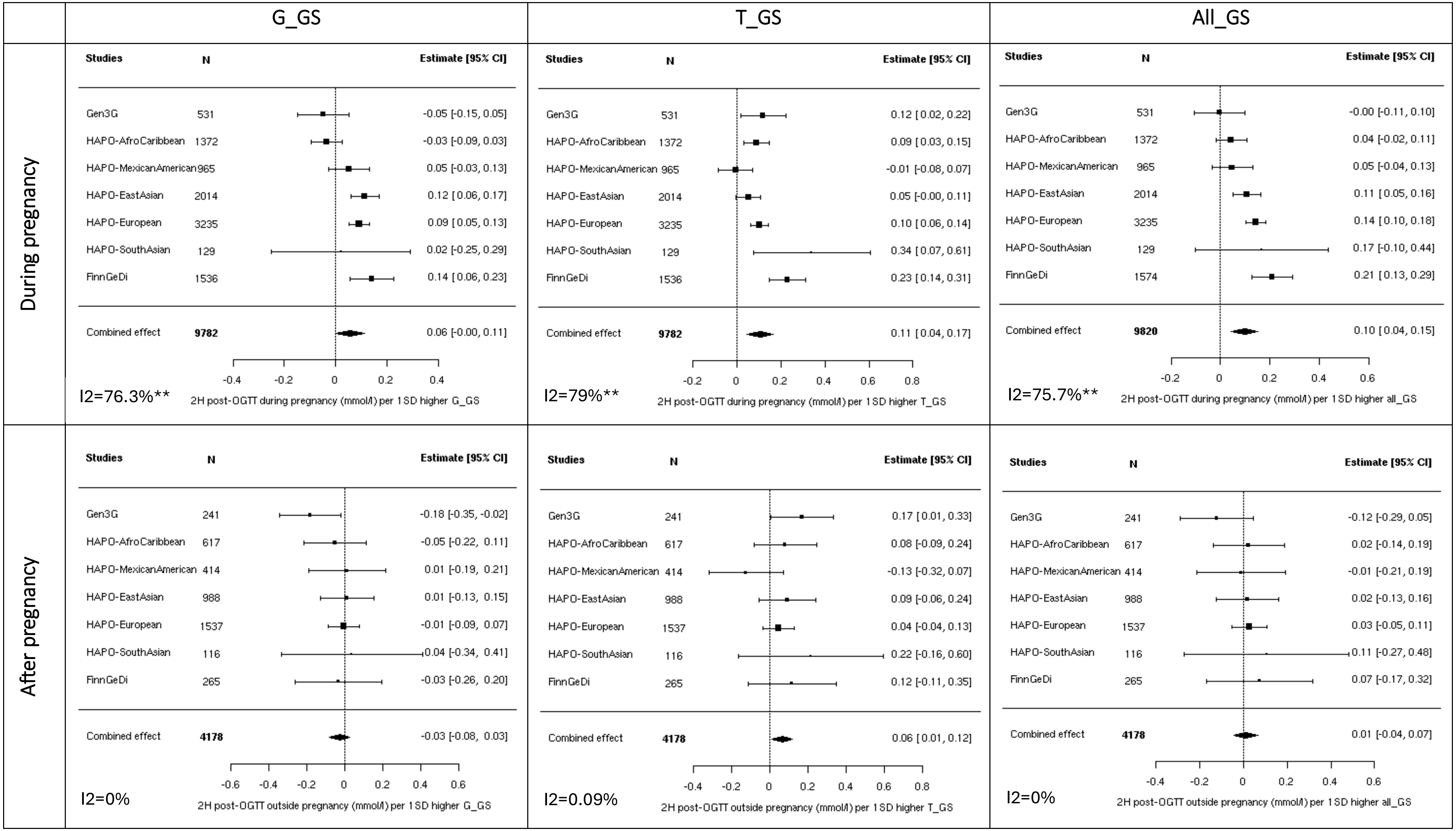


Supplementary Figure S2: Meta-analysis of the association between 2-hour glucose and genetic scores, adjusted for maternal age at glucose measurement

Analyses adjusted also for principal components and cohort-specific variables (to address unique characteristics of each dataset). Heterogeneity statistics ($I^2$) are included in the bottom left of each plot.***: p-value <.0001; **: p-value < 0.001; *: p-value < 0.05

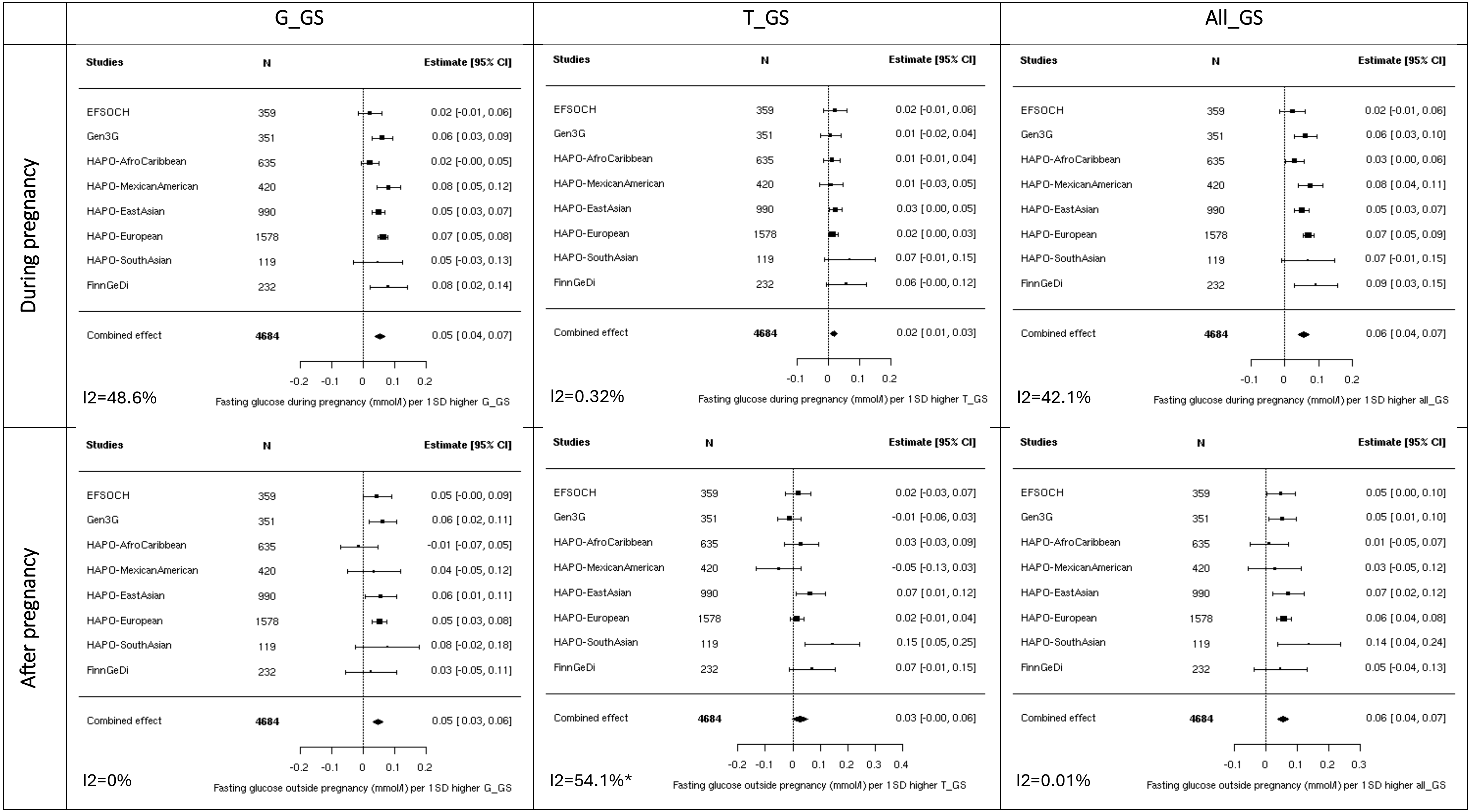


**Supplementary Figure S3: Meta-analysis of the association between fasting glucose and genetic scores, including only women who had measurements before and after pregnancy.** Analyses adjusted solely for principal components and cohort-specific variables (to address unique characteristics of each dataset). Heterogeneity statistics ($I^2$) are included in the bottom left of each plot.

***: p-value <.0001; **: p-value < 0.001; *: p-value < 0.05

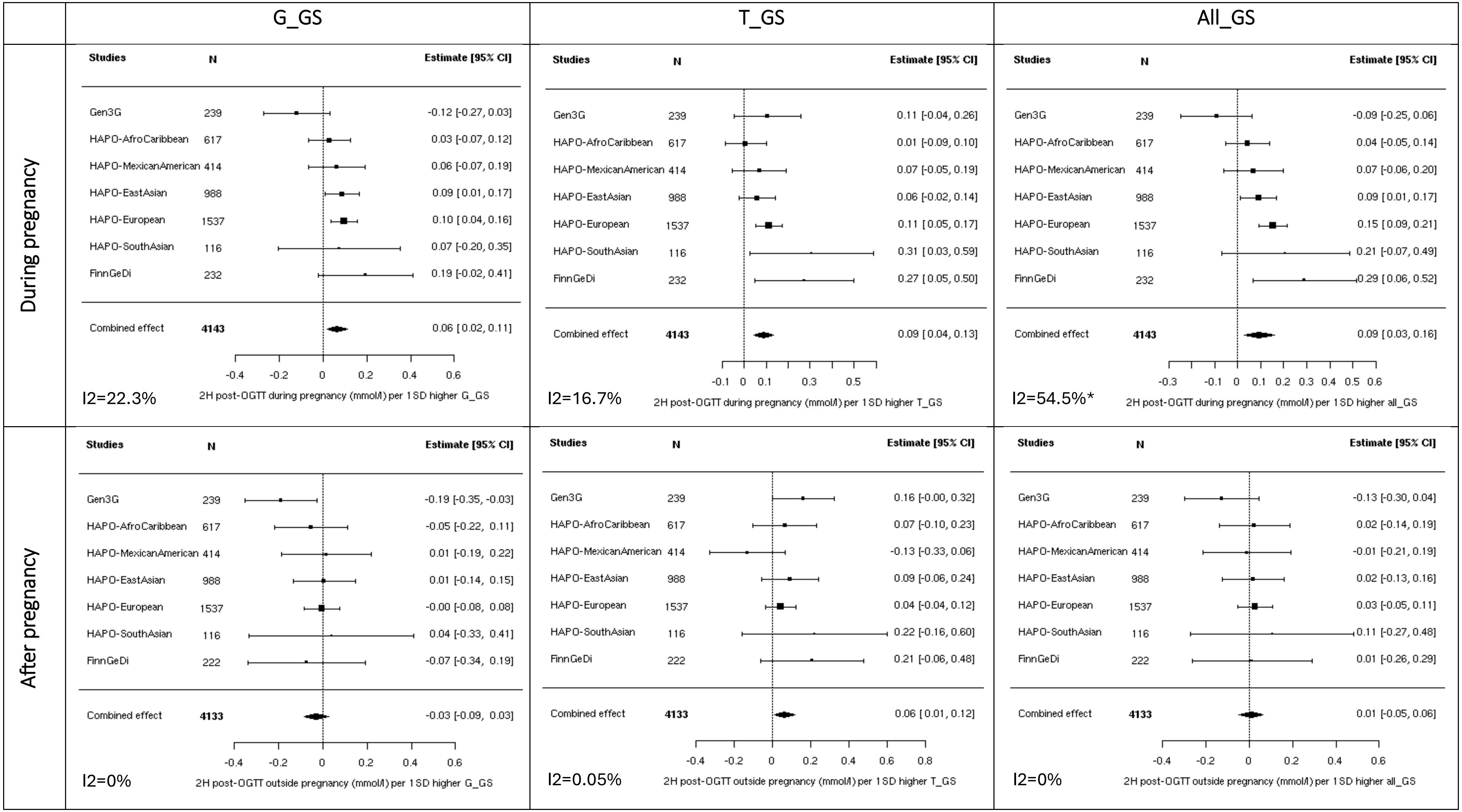


**Supplementary Figure S4: Meta-analysis of the association between 2-hour glucose and genetic scores, including only women who had measurements before and after pregnancy.** Analyses adjusted solely for principal components and cohort-specific variables (to address unique characteristics of each dataset). Heterogeneity statistics ($I^2$) are included in the bottom left of each plot.

***: p-value <.0001; **: p-value < 0.001; *: p-value < 0.05

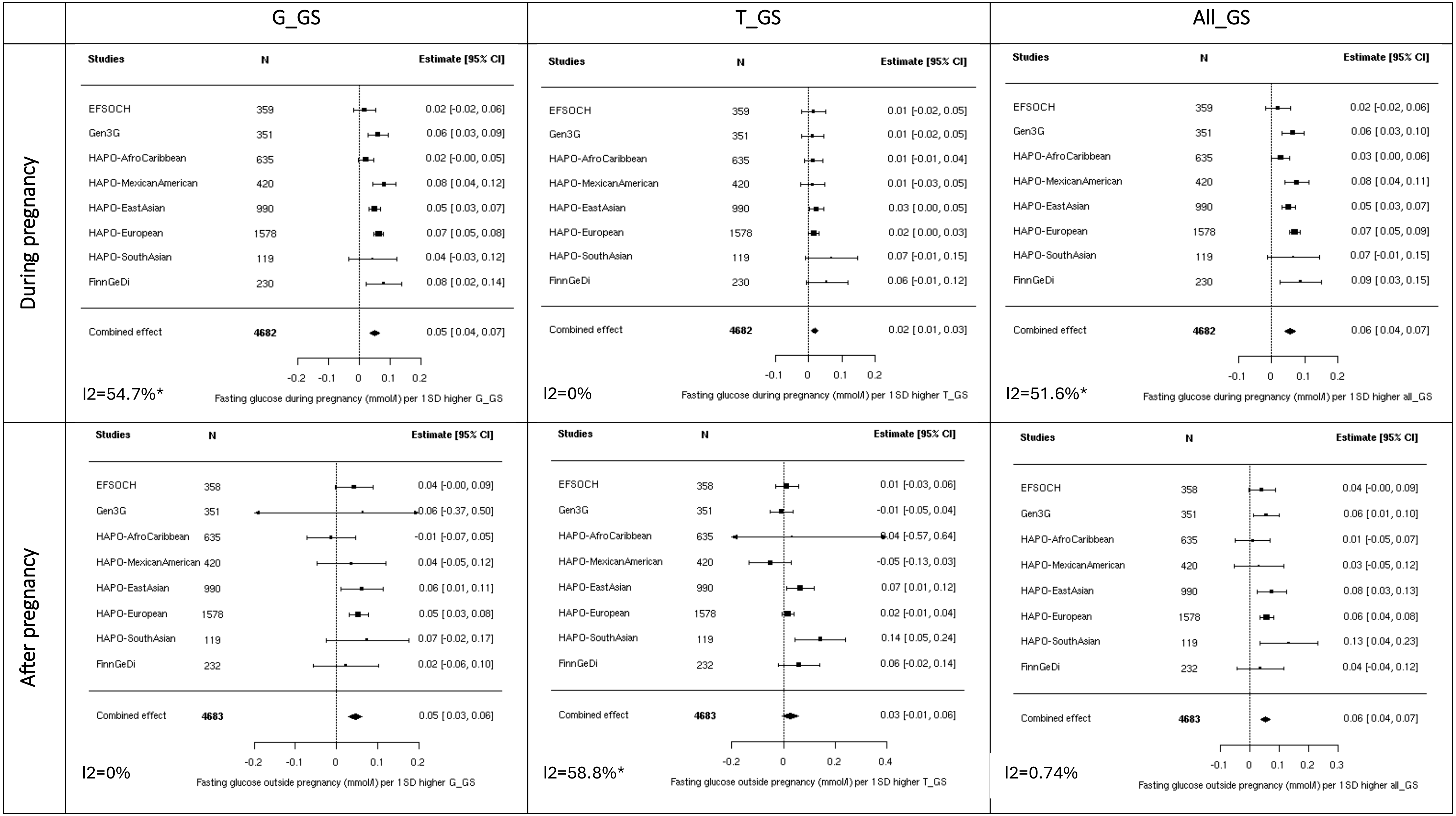


**Supplementary Figure S5: Meta-analysis of the association between fasting glucose and genetic scores, including only women who had measurements before and after pregnancy and adjusted for maternal age at glucose.** Analyses adjusted also for principal components and cohort-specific variables (to address unique characteristics of each dataset). Heterogeneity statistics ($I^2$) are included in the bottom left of each plot.

***: p-value <.0001; **: p-value < 0.001; *: p-value < 0.05

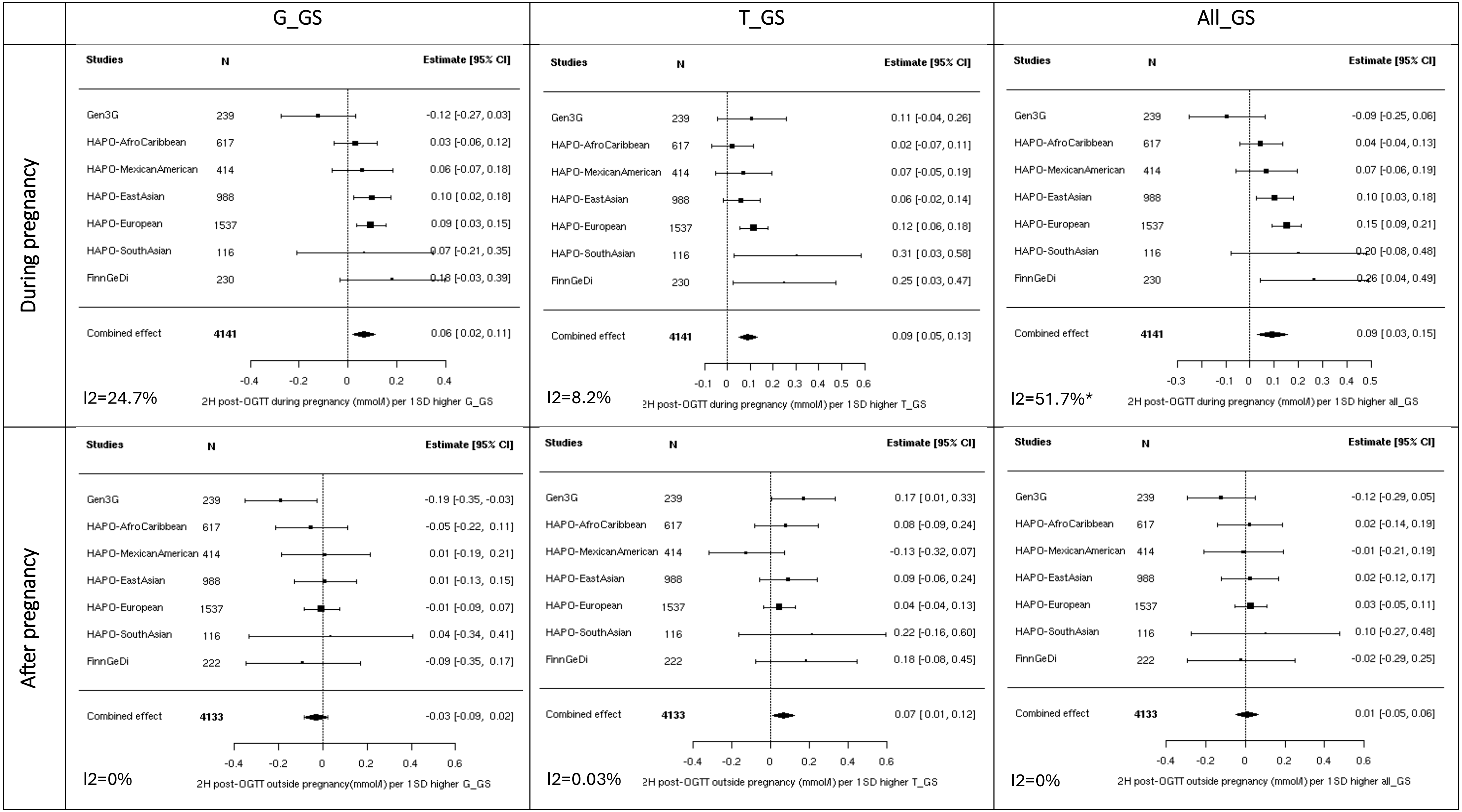


**Supplementary Figure S6: Meta-analysis of the association between 2-hour glucose and genetic scores, including only women who had measurements before and after pregnancy and adjusted for maternal age at glucose measurement.** Analyses adjusted also for principal components and cohort-specific variables (to address unique characteristics of each dataset). Heterogeneity statistics ($I^2$) are included in the bottom left of each plot.

***: p-value <.0001; **: p-value < 0.001; *: p-value < 0.05

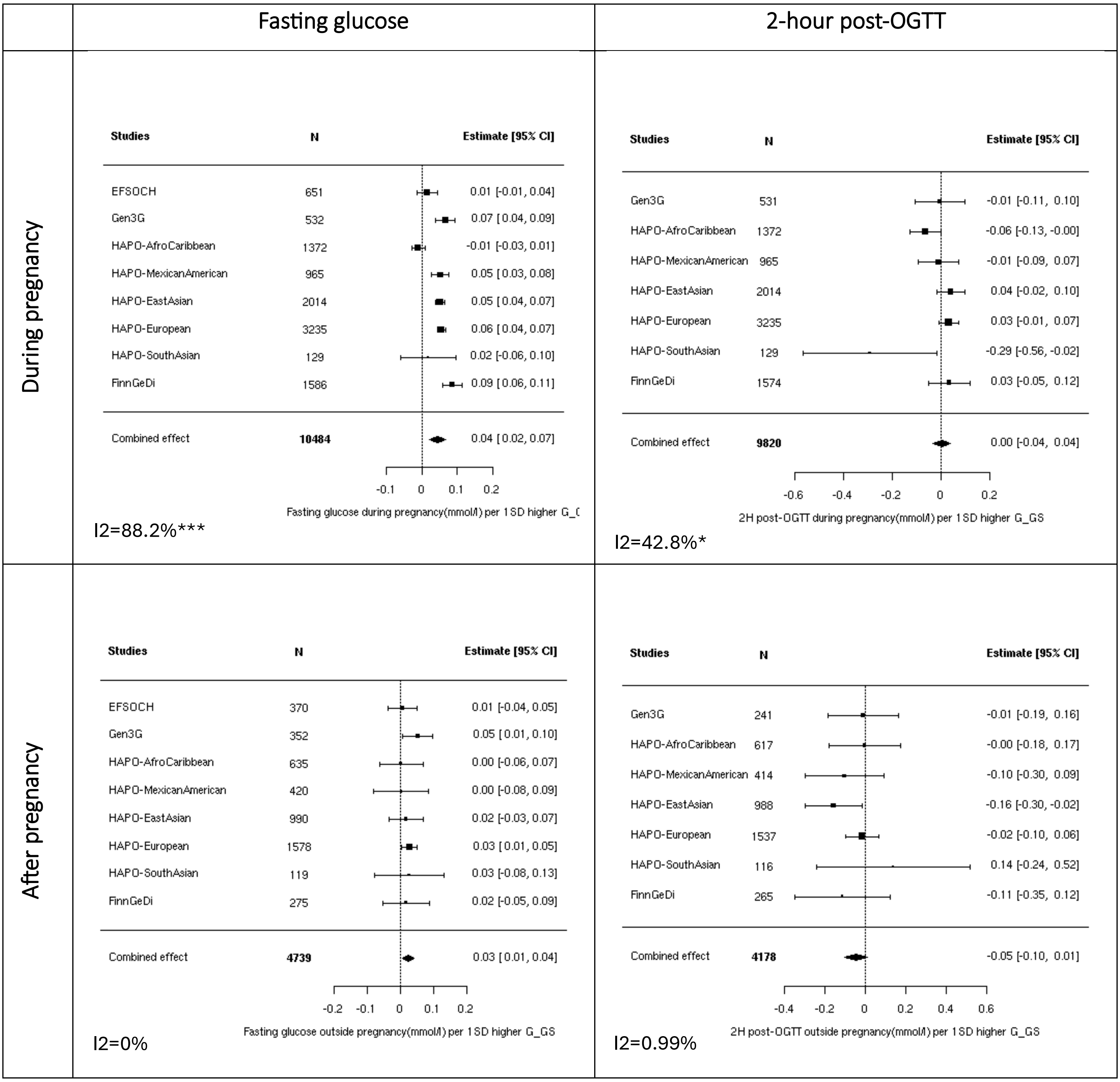


**Supplementary Figure S7: Meta-analysis of the association between glucose levels (fasting glucose and 2-hour glucose post-OGTT) and G_GS, removing *MTNR1B***

Analyses adjusted solely for principal components and cohort-specific variables (to address unique characteristics of each dataset)

***: p-value <.0001; **: p-value < 0.001; *: p-value < 0.05

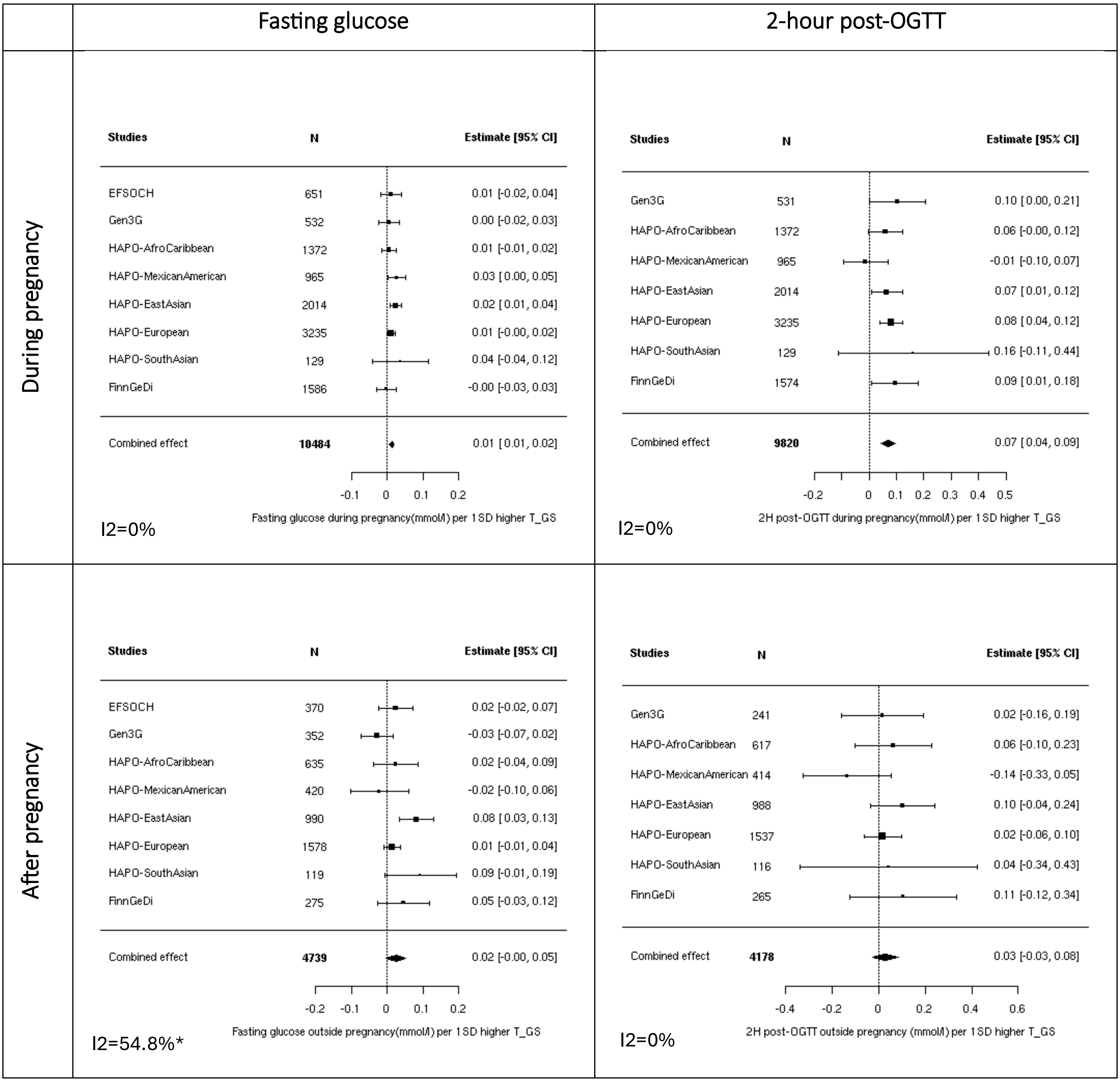


Supplementary Figure S8: Meta-analysis of the association between glucose levels (fasting glucose and 2-hour glucose post-OGTT) and T_GS, removing *TCF7L2*

* Analyses adjusted solely for principal components and cohort-specific variables (to address unique characteristics of each dataset)

***: p-value <.0001; **: p-value < 0.001; *: p-value < 0.05
